# Genomic characterization of normal and aberrant human milk production

**DOI:** 10.1101/2025.06.22.25329156

**Authors:** Yarden Golan, Sarah K. Nyquist, Zhe Liu, Dena Ennis, Jingjing Zhao, Emily Blair, Abdur Rahim Khan, Mary Prahl, Stephanie L. Gaw, Moran Yassour, Barbara E. Engelhardt, Valerie J. Flaherman, Nadav Ahituv

**Affiliations:** Division of Nutritional Sciences, Cornell University, Ithaca, USA; Gladstone Institutes, San Francisco, CA, USA; Department of Bioengineering and Therapeutic Sciences, University of California, San Francisco, USA; Institute for Human Genetics, University of California, San Francisco, California, USA; Microbiology & Molecular Genetics Department, Faculty of Medicine, The Hebrew University of Jerusalem, Israel; Department of Pediatrics, University of California, San Francisco, California, United States of America; Benioff Center for Microbiome Medicine, Division of Gastroenterology, Department of Medicine, University of California, San Francisco, CA; Division of Pediatric Infectious Diseases and Global Health, University of California, San Francisco, California, United States of America. 8Division of Maternal-Fetal Medicine, Department of Obstetrics, Gynecology, and Reproductive Sciences, University of California San Francisco, California, United States of America; Center for Reproductive Sciences, Department of Obstetrics, Gynecology, and Reproductive Sciences, University of California San Francisco, California, United States of America; The Rachel and Selim Benin School of Computer Science and Engineering, The Hebrew University of Jerusalem, Israel; Department of Biomedical Data Science, Stanford University, Stanford, CA, USA

## Abstract

Breastfeeding is essential for reducing infant morbidity and mortality, yet exclusive breastfeeding rates remain low, often due to insufficient milk supply. The molecular causes of low milk production are not well understood. Fresh milk samples from 30 lactating individuals, classified by milk production levels across postpartum stages, were analyzed using genomic and microbiome techniques. Bulk RNA sequencing of milk fat globules (MFG), milk cells, and breast tissue revealed that MFG-derived RNA closely mirrors luminal milk cells. Transcriptomic and single-cell RNA analyses identified changes in gene expression and cellular composition, highlighting key genes (GLP1R, PLIN4, KLF10) and cell-type differences between low and high producers. Infant microbiome diversity was influenced by feeding type but not maternal milk supply. This study provides a comprehensive human milk transcriptomic catalog and highlights that MFG could serve as a useful biomarker for milk transcriptome analysis, offering insights into the genetic factors influencing milk production.

## Introduction

Breastfeeding is associated with reduced risk for morbidity and mortality for the infant early and later in life(*1*). The World Health Organization (WHO) recommends exclusively breastfeeding for the first six months of life, and a continuation of breastfeeding for up to two years or longer with the introduction of complimentary foods(*2*). However, the rates and duration of exclusive breastfeeding are low, with 48% of infants worldwide(*3*) and 24% of infants in the US(*4*) being exclusively breastfed at six months of age. One of the main reasons for early weaning, reported by approximately 35% of individuals that wean their infants earlier than recommended, is perceived insufficient milk supply (PIMS)(*5*). In some cases, early intervention and support for the mother can improve milk production and help to continue breastfeeding(*6*). However, little is known about the molecular mechanism leading to PIMS and about the molecular changes in the mammary gland in cases of perceived or measured low milk production. This gap in knowledge lead to very limited treatment options in these cases (*7*, *8*). Previous studies found an association between maternal obesity and low milk production(*9*, *10*). Overweight and obese women were less likely to initiate and maintain breastfeeding and more likely to report that their infant is not satisfied with breast milk alone, in comparison to women with normal weight(*11*, *12*). In addition, high levels of systemic TNF-alpha and inflammation were suggested to be associated with low milk production in obesity(*13*, *14*), as well as insulin dysregulation(*15*). Nevertheless, PIMS and low milk production is also common among women who are not overweight(*5*, *16*). Larger and controlled clinical studies are needed to better understand these relationships and to examine improved intervention protocols.

On the opposite end of the milk production spectrum, some women suffer from over production (hyperlactation) that may be idiopathic or may be caused by over pumping or use of galactagogues (i.e., substances thought to increase the rate of human milk synthesis but are not approved by the FDA)(*17*). Hyperlactation is also at risk for early weaning, due to higher frequencies of breast pain, plugged ducts, and mastitis(*17*). Global characterization of the cellular signaling, genes, regulatory elements, and pathways associated with low or high milk production may help to better diagnose, treat, and support these individuals.

Genetic studies on human breastfeeding complications are sparse in part because these conditions are not well documented in medical records, and we lack a gold standard method to diagnose hypo or hyperlactation. Almost all genome-wide association studies (GWAS) on milk yield have been performed in cows and other dairy animals, finding associations between single nucleotide polymorphisms (SNPs) and milk production traits(*18*). One example is a SNP associated with the gene *DGAT1*, a key enzyme that catalyzes the final step of triglyceride synthesis in mammary gland cells, and is associated with milk yield and fat and lactose production traits(*19–23*). To date, only one human study has been published showing a single genetic variant associated with milk production and breastfeeding duration(*24*). In this candidate gene study, a single SNP in the milk fat globule EGF and factor V/VIII domain containing gene (*MFGE8*) was found to be associated with PIMS, breastfeeding exclusivity, and duration. In addition, there are a few studies on the relationship between human maternal genetics and milk composition, which have focused on human milk oligosaccharides (HMOs), zinc transport, and fatty acids(*25–31*). While these studies have not tested for associations with milk production, they nevertheless provide support for the role of various genes in the milk production process to develop more effective diagnostics and interventions. Moreover, one previous study on milk gene expression found higher expression of the *TARDBP* gene in exclusive breastfeeding individuals compared to those supplemented with formula 3-5 days postpartum, often an indication of issues with milk production (*32*). All together, these studies suggest that human milk production or milk yield might also be regulated at the gene expression level.

Recent work used functional genomics tools to characterize the human milk transcriptome(*33–35*) and have found that milk fat globules (MFG) can serve as an information-rich, non-invasive biospecimen for learning about mammary gland function(*33–37*). MFG contains high amounts of RNA that is easy to extract compared to the relatively low amount of RNA extracted from milk cells, and it does not require sorting for specific cell populations as they are secreted from epithelial cells (*38*, *39*). During this process, MFG are coated by the epithelial cell membrane, and some of the cell’s cytosol is secreted into the milk in the crescent structure between the MFG membrane and the cell membrane(*40*). However, to date, no direct comparison between MFG RNA and human milk cells was published, and it is unclear if specific RNA is targeted to be secreted in these compartments. In addition, we and others used single-cell RNA sequencing (scRNA-seq) to characterize various milk cell type populations, at different breastfeeding time points, finding 2-6 different lactocyte cell type populations(*41–46*) and cross-talk between these cell types and immune cells(*44*). The two main epithelial cell subtypes that are found in milk are referred to as luminal cells 1 and 2 (LC1s and LC2s) (*42–45*). It is unclear whether these epithelial subtypes play different roles in milk production or MFG secretion. A better understanding of the MFG transcriptome and the cells that produce them could dramatically enhance studies that use these accessible milk fractions as a proxy biomarker to understand the mammary gland function during lactation.

Here, we used RNA-seq and scRNA-seq to better understand the human milk transcriptome under normal and aberrant milk production conditions. We show that the MFG transcriptome resembles milk luminal cells with more similarity to the LC2 cells, and can be used as a milk biomarker studying the function of the mammary gland luminal cells during lactation. We also provide an important database of genes that are upregulated during lactation compared to non-lactating breast tissue. Using samples from individuals that differ in their milk production, stratified into low, normal, and high production groups, we identify changes in the cellular transcriptome and milk cell type composition between these groups. In addition, we tested whether maternal low milk production impacts the infant microbiome, and whether infant formula supplementation, which occurs more frequently in infants nursing from mothers with low milk production, affects the infant microbiome. Taken together, our findings shed light on molecular and cellular changes under different levels of milk production.

## Results

### Collection of human milk samples with differences in milk production

To identify the molecular factors associated with milk production, we collected fresh milk samples from 30 lactating individuals during various lactation stages (**Fig. 1A**). Participants included nine individuals with low milk production, seven individuals with high milk production, all referred by lactation consultants after a breast exam and infant latching examination (further details in **Table S1**), and 14 individuals who self-reported normal milk production (recruited from social media and distributed flyers at UCSF clinics). In addition to breastfeeding consultants’ categorization (**Table S1**), mothers also reported perception of their milk production, which differed across the study groups (chi-square p ≤ 0.001; **Table 1**). One mother who self-reported low milk production but was categorized by the lactation consultant as a normal supplier was excluded from further analysis. Furthermore, we assessed maternal reports about their infants’ satisfaction with the amount of breast milk the infants received (“Is the infant satisfied with mother’s own milk?”); we found this measure to also differ across the study milk-production groups (chi-square p ≤ 0.001; **Table 1**). Mothers with low milk production were more likely to supplement their infant’s diet with baby formula or donor milk; 7 out of the 9 mothers supplemented with infant formula in the first 8 days postpartum (**Fig. 1**; additional information on formula supplementation and lactation consultant summary for each participant in **Table S1**). Using data collected at enrollment, we found that individuals with normal milk production tended to report that they were breastfed as an infant, compared to the low and high groups that report less-frequently that they were breastfed as infants (ANOVA p ≤ 0.042; **Table 1**). In addition, there was a trend of self-report of delay in the day they felt that the milk “came in” (corresponding to lactogenesis II/secretory activation) in the low production group (ANOVA p ≤ 0.072; **Table 1**), which was previously shown to be associated with unintended breastfeeding reduction and cessation(*47*). Maternal BMI did not differ significantly between the groups in our cohort (ANOVA, p > 0.1; **Table 1**). Participants with low, normal, and high milk production showed relevant differences between perceived milk production, but no difference in other known risk factors for low milk production, including maternal age, and delivery mode (**Table 1**). Each individual contributed a mean of 2 ± 1.6 samples to the study.

**Fig. 1.**
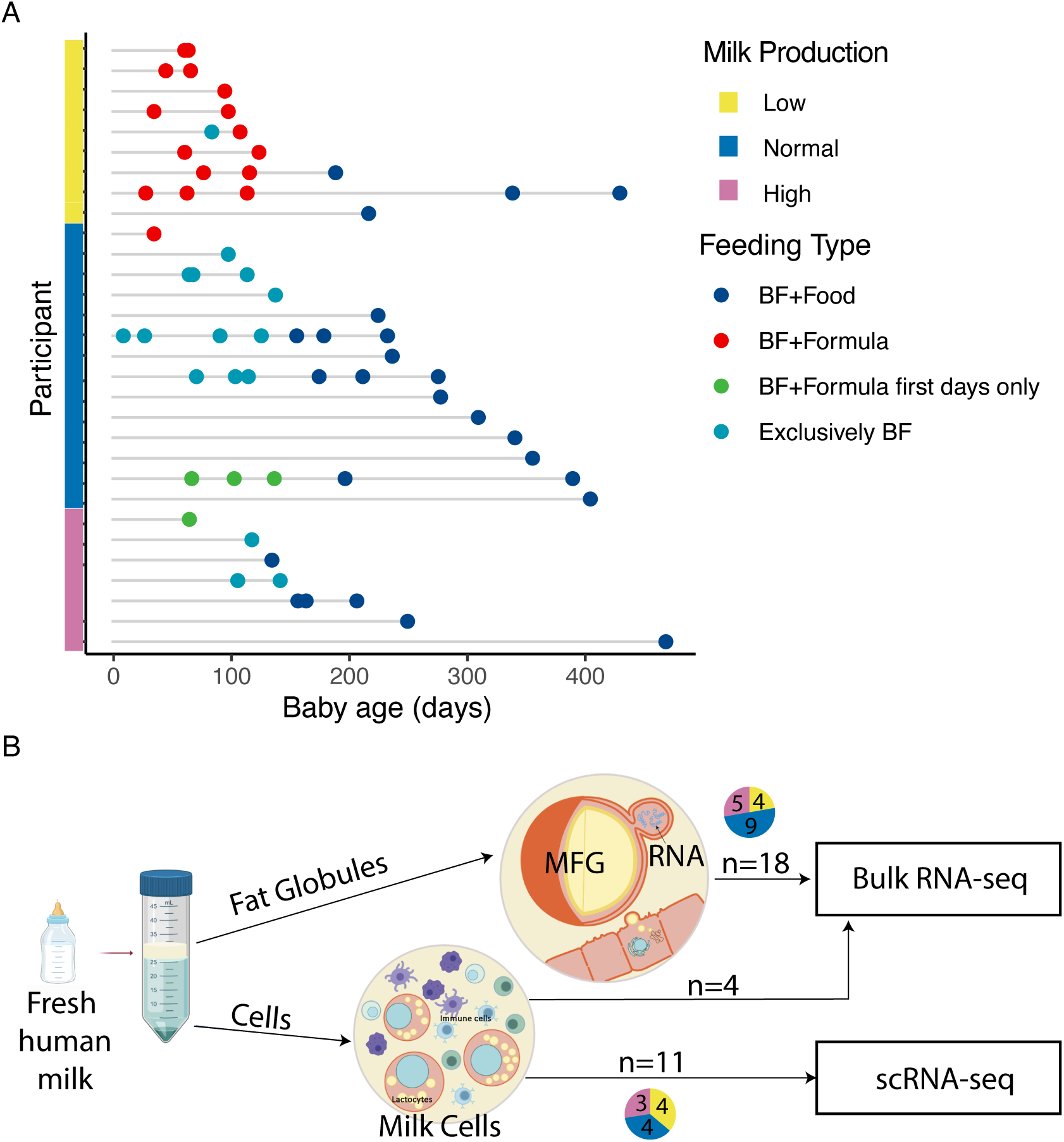
Samples and milk fractions used in this study. (A) Graph showing the samples used in the study, their categorization into milk production groups, lactation stage, and infant feeding type (BF = breastfeeding) at the time of sample collection. (B) Schematic showing the milk fraction observed after centrifugation of fresh milk samples (milk fat globules (MFG) at the top and milk cells at the bottom) and the experiments carried out on them.

**Table 1:** study participants characteristics.

| Category | Overall | Low | Normal | High | p value<sup>#</sup> | Test |
| --- | --- | --- | --- | --- | --- | --- |
| **n** | **30** | **9** | **14** | **7** |  |  |
| **Age (years) (mean (SD))** | 35.85 (3.26) | 36.45 (3.15) | 35.93 (3.08) | 34.93 (4.00) | ns | ANOVA |
| **Ethnic group (%)** |  |  |  |  | ns | Chi-square |
| **White** | 20 (66.7) | 7 ( 77.8) | 9 ( 64.3) | 4 ( 57.1) |  |  |
| **Asian Indian** | 1 ( 3.3) | 0 ( 0.0) | 1 ( 7.1) | 0 ( 0.0) |  |  |
| **Chinese** | 3 (10.0) | 1 ( 11.1) | 1 ( 7.1) | 1 ( 14.3) |  |  |
| **Hispanic** | 2 ( 6.7) | 0 ( 0.0) | 1 ( 7.1) | 1 ( 14.3) |  |  |
| **Other** | 3 (10.0) | 1 ( 11.1) | 1 ( 7.1) | 1 ( 14.3) |  |  |
| **Did not answer** | 1 ( 3.3) | 0 ( 0.0) | 1 ( 7.1) | 0 ( 0.0) |  |  |
| **BMI (mean (SD))** | 24.37 (4.06) | 26.58 (4.02) | 22.94 (2.51) | 24.40 (5.97) | ns | ANOVA |
| **Number of pregnancies (mean (SD))** | 2.22 (1.00) | 2.00 (1.07) | 2.00 (0.71) | 2.83 (1.17) | ns | ANOVA |
| **Number of live birth (mean (SD))** | 1.50 (0.58) | 1.12 (0.35) | 1.57 (0.65) | 1.83 (0.41) | † | ANOVA |
| **Age at first pregnancy (mean (SD))** | 33.48 (2.74) | 34.75 (1.91) | 33.44 (2.40) | 31.83 (3.60) | ns | ANOVA |
| **Breastfeeding experience (%)** |  |  |  |  | ns | Chi-square |
| **Don't have other kids** | 13 (44.8) | 7 ( 77.8) | 7 ( 50.0) | 1 ( 16.7) |  |  |
| **Breastfed 1 child before** | 1 ( 3.4) | 2 ( 22.2) | 6 ( 42.9) | 5 ( 83.3) |  |  |
| **Breastfed 2 children before** | 15 (51.7) | 0 ( 0.0) | 1 ( 7.1) | 0 ( 0.0) |  |  |
| **Weight before pregnancy (kilograms) (mean (SD))** | 63.74 (12.07) | 69.92 (14.72) | 60.71 (7.62) | 61.86 (14.39) | ns | ANOVA |
| **Breastfed as a child = Yes (%)** | 21 (72.4) | 5 ( 55.6) | 13 ( 92.9) | 3 ( 50.0) | * | Chi-square |
| **Age of first menstrual (mean (SD))** | 13.09 (1.38) | 13.50 (1.60) | 13.00 (1.00) | 12.67 (1.63) | ns | ANOVA |
| **Baby gender = Male (%)** | 17 (58.6) | 5 ( 62.5) | 8 ( 57.1) | 4 ( 57.1) | ns | Chi-square |
| **Gestational age at delivery (mean (SD))** | 39.10 (1.27) | 39.33 (1.41) | 39.29 (1.27) | 38.43 (0.98) | ns | ANOVA |
| **Mode of delivery (%)** |  |  |  |  | ns | Chi-square |
| **Vaginal** | 27 (90.0) | 8 ( 88.9) | 14 (100.0) | 5 ( 71.4) |  |  |
| **C-section after failed trail of labor** | 2 ( 6.7) | 1 ( 11.1) | 0 ( 0.0) | 1 ( 14.3) |  |  |
| **Intended C-section** | 1 ( 3.3) | 0 ( 0.0) | 0 ( 0.0) | 1 ( 14.3) |  |  |
| **Day post partum that milk came in (Lactogenesis stage II) (mean (SD))** | 3.45 (1.75) | 4.62 (2.26) | 3.04 (1.48) | 2.86 (0.90) | † | ANOVA |
| **First try breastfeeding (%)** |  |  |  |  | ns | Chi-square |
| **Immediately** | 14 (46.7) | 2 ( 22.2) | 7 ( 50.0) | 5 ( 71.4) |  |  |
| **< 30 minutes** | 9 (30.0) | 4 ( 44.4) | 4 ( 28.6) | 1 ( 14.3) |  |  |
| **30 min - 1 hour after birth** | 4 (13.3) | 2 ( 22.2) | 1 ( 7.1) | 1 ( 14.3) |  |  |
| **2 - 6 hours after birth** | 1 ( 3.3) | 0 ( 0.0) | 1 ( 7.1) | 0 ( 0.0) |  |  |
| **>24 hours after birth** | 2 ( 6.7) | 1 ( 11.1) | 1 ( 7.1) | 0 ( 0.0) |  |  |
| **Milk production assessment (%)** |  |  |  |  | *** | Chi-square |
| **I feel that I produce not enough breast milk to meet my baby's needs.** | 9 (30.0) | 9 (100.0) | 0 ( 0.0) | 0 ( 0.0) |  |  |
| **I feel that I produce normal levels of breast milk to meet my baby's needs.** | 14 (46.7) | 0 ( 0.0) | 14 (100.0) | 0 ( 0.0) |  |  |
| **I feel that I produce too much breast milk to meet my baby's needs.** | 7 (23.3) | 0 ( 0.0) | 0 ( 0.0) | 7 (100.0) |  |  |
| **My infant is satisfied with my own milk. (%)** |  |  |  |  | *** | Chi-square |
| **Agree strongly** | 16 (53.3) | 0 ( 0.0) | 10 ( 71.4) | 6 ( 85.7) |  |  |
| **Agree moderately** | 3 (10.0) | 0 ( 0.0) | 3 ( 21.4) | 0 ( 0.0) |  |  |
| **Agree slightly** | 2 ( 6.7) | 1 ( 11.1) | 1 ( 7.1) | 0 ( 0.0) |  |  |
| **Disagree moderately** | 3 (10.0) | 3 ( 33.3) | 0 ( 0.0) | 0 ( 0.0) |  |  |
| **Disagree strongly** | 5 (16.7) | 5 ( 55.6) | 0 ( 0.0) | 0 ( 0.0) |  |  |
| **Neither agree nor disagree** | 1 ( 3.3) | 0 ( 0.0) | 0 ( 0.0) | 1 ( 14.3) |  |  |

### The MFG RNA signature recapitulates the lactocyte transcriptome

MFG RNA is readily accessible, and contains high amounts of RNA compared to the cell pellet in each milk sample. However, the origin of this RNA and direct comparison to milk cells RNA from the same samples was never performed, leaving a gap in knowledge about the usability of MFG RNA as a biomarker for the different cell types in the mammary gland. To address this, bulk RNA-seq was performed on a subset of four paired human milk cells and MFG samples obtained from the same milk sample of three individuals (days 139-481 postpartum), followed by transcriptomic comparisons (sample information in **Table 2**). We found 1,696 genes to be differentially expressed (DE) between the milk cells and MFG; of those, 1,454 genes showed higher expression in human milk cells and only 242 genes showed higher expression in MFG (**Fig. 2A**). We performed gene ontology (GO) enrichment analysis using clusterProfiler (adjusted p-values calculated using Benjamini-Hochberg (BH) correction(*48*)). We found that genes with higher expression in milk cells are associated with GO terms related to *leukocyte and lymphocyte differentiation* and *cytokines*, capturing the presence versus absence of immune cells in milk cells versus the MFG fraction, respectively (**Fig. 2B**). For the genes with higher expression in MFG, we observed no significant GO enrichment. The overexpressed genes in the MFG fraction overlapped several GO term lists (adjusted p values > 0.05) including *positive regulation of lipid localization*, the *regulation of fatty acid transport,* and the *acetyl-CoA metabolic process* (**Fig. 2C**). This analysis showed that most of the MFG transcriptome is also present in the milk cells fraction. In contrast, if one is interested in immune function during lactation, the cell pellet should be used, not MFG RNA.

**Fig. 2:**
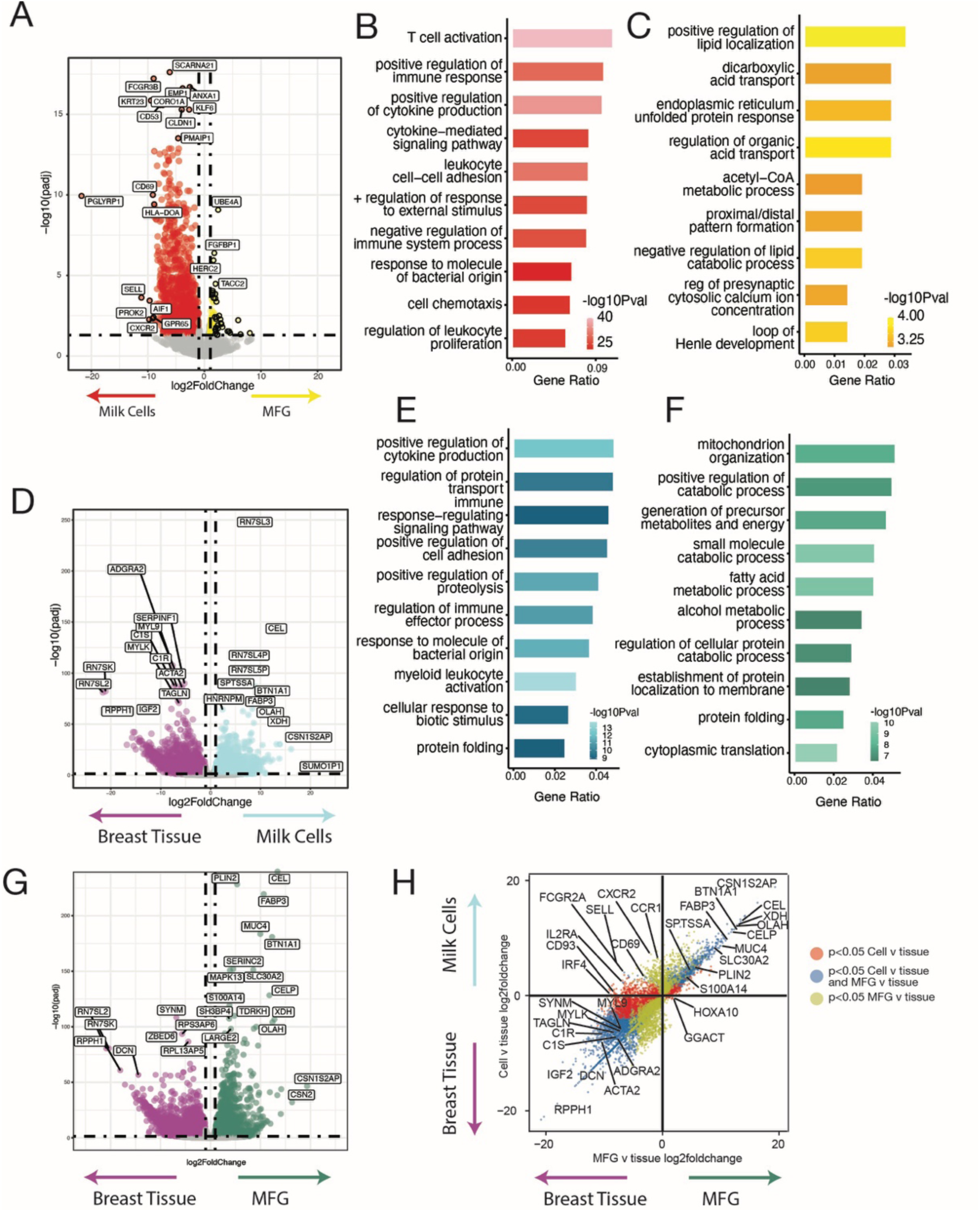
Transcriptomic comparison between MFG, milk cells and non-lactating mammary gland tissue. (A) Volcano plot showing RNA-seq differentially expressed (DE) genes between milk cells (*left side*) and MFG (*right side*) as detected by DESeq2(*51*). A log-fold change >1 cutoff and BH adjusted p-value<0.05 was used. (B-C) Gene Ontology Biological Process (GOBP) pathway analysis(*48*) of DE genes from (A) including pathways upregulated in milk cells (B) and in MFG (C). (D) Volcano plot of DE genes comparing milk cells (*left side*) and non-lactational breast tissue (*right side).* (E) Corresponding GOBP pathway analysis of DE genes up in milk cells from (D) (*right side*). (F) Volcano plot of DE genes between MFG (*left side*) and non-lactational breast tissue (*right side*). (G) Corresponding GOBP pathway analysis of DE genes up in MFG from (F) (*right side*). (H) Dot plot of the fold change of the DE genes from (D) and (G) showing similarity in gene expression between milk cells and MFG compared to breast tissue (blue) and DE genes in cells (*red*) or MFG (*green*). Full gene and GOBP lists in **Table S2**. Gene ratio is the ratio of input genes that are annotated in a term (pathway of genes).

**Table 2:** Samples sequences for different analysis in the study. “y” represents samples that were sequenced in the different assays performed in this study. Samples that were not included in the microbiome analysis due to late time postpartum or low number of participants from high production group, are marked with *.

| Participant ID | Milk supply group | Days postpartum | MFG RNA-seq | Cell bulk RNA-seq | Cell scRNA-seq | Infant stool microbiome | maternal stool microbiome |
| --- | --- | --- | --- | --- | --- | --- | --- |
| ID1024 | High | 208 | y |  |  |  |  |
| ID1025 | High | 106 | y |  | y | y* | y* |
| ID1026 | High | 84 | y |  | y |  |  |
| ID1012 | High | 469 | y | y |  |  |  |
| ID1012 | High | 481 | y | y |  |  |  |
| ID1024 | High | 158 |  |  | y | y* | y* |
| ID1018 | Low | 46 |  |  |  | y | y |
| ID1018 | Low | 67 | y |  |  | y |  |
| ID1023 | Low | 28 |  |  |  | y | y |
| ID1023 | Low | 64 | y |  | y | y | y |
| ID1027 | Low | 95 |  |  |  | y | y |
| ID1028 | Low | 85 | y |  | y | y | y |
| ID1028 | Low | 106 |  |  |  | y |  |
| ID1029 | Low | 62 | y |  | y |  |  |
| ID1030 | Low | 36 |  |  |  | y |  |
| ID1030 | Low | 99 |  |  | y | y | y |
| ID1036 | Low | 62 |  |  |  | y | y |
| ID1036 | Low | 65 |  |  |  | y | y |
| ID1037 | Low | 84 |  |  |  | y | y |
| ID1016 | Normal | 68 | y |  | y | y | y |
| ID1016 | Normal | 94 | y |  |  | y | y |
| ID1016 | Normal | 138 |  |  |  | y | y |
| ID1020 | Normal | 70 | y |  | y | y | y |
| ID1020 | Normal | 94 | y |  |  | y | y |
| ID1020 | Normal | 116 | y |  |  | y | y |
| ID1020 | Normal | 213 | y |  |  | y* | y* |
| VPL131 | Normal | 68 | y |  |  |  |  |
| ID1013 | Normal | 386 | y | y |  |  |  |
| ID1014 | Normal | 139 | y | y |  |  |  |
| ID1020 | Normal | 165 |  |  | y |  |  |
| ID1033 | Normal | 61 |  |  | y | y | y |
| ID1033 | Normal | 114 |  |  |  | y |  |

MFGs, secreted from mature and active milk-producing cells, primarily contain milk protein-related transcripts(*33*). To better understand aberrant lactation, our first goal was to identify genes and pathways upregulated during normal lactation. We compared MFG and milk cell transcriptomes to the non-lactating mammary gland transcriptome to determine whether similar pathways are upregulated in both, providing insight into the representativeness of MFG RNA as a model for the milk-producing cells’ transcriptome. For this analysis, we compared publicly available bulk RNA-seq data from non-lactational breast tissue(*49*) to bulk RNA-seq from milk cells and MFG in our samples. We identified 11,810 DE genes between non-lactational breast tissue and milk cell or MFG fractions (**Table S2**). Comparing the milk cell transcriptome to non-lactational breast tissue, we found 3,462 genes to be highly expressed in milk cells, many of them known to be related to milk production (including the *casein* family genes), and 5,796 genes highly expressed in non-lactational breast tissue compared to milk cells (**Fig. 2D, Table S2**). The genes upregulated in milk cells were enriched for pathways associated with *myeloid leukocyte activation* (BH adjusted p ≤ 9.8e-11) and *positive regulation of cytokine production* (BH adjusted p ≤ 1.3e-09), as well as *regulation of protein transport* (BH adjusted p ≤ 1.4e-07) capturing the higher activity of immune cells during lactation than in non-lactational breast tissue (*50*)(**Fig. 2E, Table S2**). The upregulated pathways in the MFG transcriptome compared to the breast tissue transcriptome were associated with *cytoplasmic translation* (BH adjusted p ≤ 5.3e-07), *small molecule catabolic processes* (BH adjusted p ≤ 1.1e-06), and *fatty acids metabolic process* (BH adjusted p ≤ 1.4e-06), which are all activated during lactation (**Fig. 2F, Table S2**). Comparing the MFG transcriptome to non-lactational breast tissue, we found 2,935 genes to be highly expressed in MFG and 6,122 genes highly expressed in non-lactational breast tissue compared to MFG (**Fig. 2G, Table S2**). Moreover, we found a large overlap in DE genes (DESeq2(*51*) adjusted p ≤ 0.05) between milk cells and MFG compared to non-lactational breast tissue, including known milk production genes such as *CEL, OLAH, SPP1,* and *CSN3,* with 2,148 jointly upregulated in milk cells and MFG, and 4,177 genes jointly downregulated versus non-lactational breast tissue (**Fig. 2H, Table S2**). In summary, our results show that the MFG transcriptome closely resembles milk cells and may be used as an accessible and homogenous proxy marker to analyze epithelial cell gene expression during lactation. This analysis also provides an important database for gens that are upregulating during lactation and can facilitate future studies focusing on lactation specific pathways.

### The MFG transcriptome is more similar to the lactocyte LC2 subtype compare to LC1

Previous studies have identified two main epithelial subtypes in human breast milk—luminal cells 1 and 2 (LC1s and LC2s) (*42–45*). We next set out to characterize whether the MFG RNA is more similar to one of these subtypes, in order to better understand the origin of MFG RNA and the specific roles of these sub cell-types in milk production.

To achieve this, we analyzed scRNA-seq data generated using the 10X Genomics platform from 11 milk cells samples in our cohort (**Fig. 1B**, **Table 2, Table S3**). After filtering, we had 17,000 cells that were clustered into nine broad cell type categories matching previous milk scRNA-seq reports(*42–45*). These cell type categories included: i) six immune cell clusters consisting of T cells, natural killer cells, B cells, plasma cells, dendritic cells (DC), and macrophages; ii) two large luminal epithelial cell clusters LC1 and LC2; and iii) a small dividing epithelial cell cluster (**Figures 3A and S1**, **Table 3**). We then used the BisqueRNA package(*52*), which uses scRNA-seq signatures to deconvolve bulk transcriptomes into component cell type proportions as a proxy for cell type of origin for the MFG transcripts. First, we generated a reference signature of general single cell types found in our scRNA-seq samples (**Fig. 3A**; LC1s, LC2s, macrophages and dendritic cells, B cells, T cells, NK cells, and plasma cells). Then, we applied this reference signature to identify which cell population is more highly represented in the MFG bulk RNA-seq data. We found that LC2 cells had the highest proportion in 12 out of the 14 MFG bulk RNA-seq samples that we examined (**Fig. 3B**). In the two samples for which LC2 did not have the highest proportion, the macrophage and DC signatures had the highest proportions and LC1 and LC2 had the second highest proportions.

**Fig. 3:**
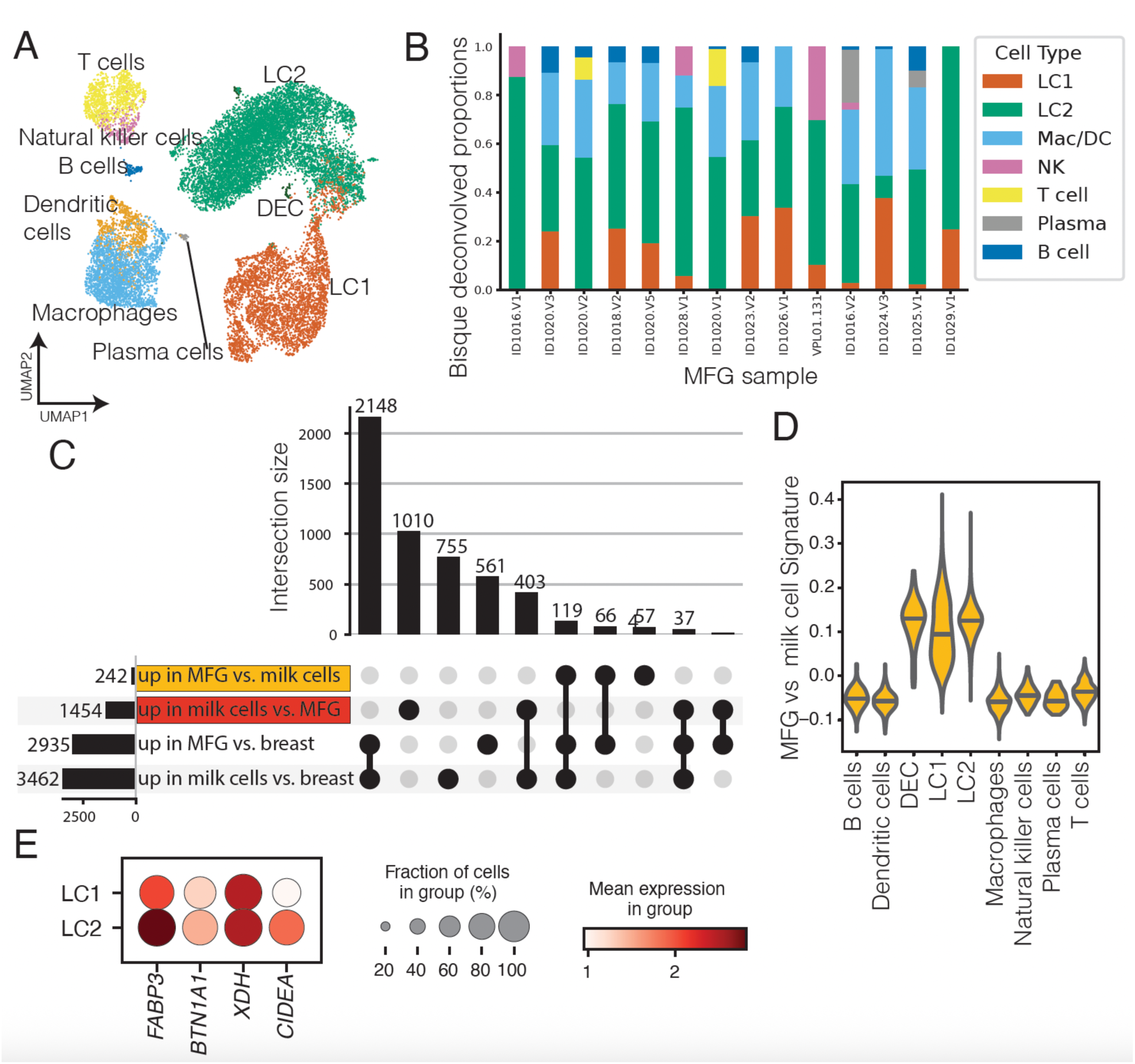
The MFG transcriptome is more similar to the epithelial subcluster LC2 compare to LC1. (A) UMAP low dimensional visualization of scRNA-seq data colored by cell type cluster. (B) BisqueRNA package (52) was used to deconvolve bulk transcriptomes from MFG into component cell type proportions as a proxy for cell type. Cells proportion as represent in MFG RNA transcript (y-axis) was calculated for each participant (x-axis). (**C**) Bar chart showing the numbers and the overlap of DE genes between milk cells, MFG, and non-lactational breast tissue. Upper bars indicate intersection size between sets indicated by dark dots below, left horizontal bars indicate size of each set of genes. (D) Violin plot of the gene set signature score (see Methods), calculated from genes upregulated in MFG compared to milk cells in bulk RNA sequencing (highlighted in yellow in (B)). The score is plotted for each cell across the cell types identified in scRNA-seq data (x-axis). (E) Genes associated with MFG formation in the LC1 and LC2 epithelial clusters. Dot size represents the percentage of cells in cluster (y-axis) expressing the gene at a level > 0 and color indicates the mean log2-normalized expression of that gene in the cells of that cluster.

**Table 3:** scRNA-seq cell type proportions per human milk sample.

| Cell type | Low Supply |  | Normal Supply |  | High Supply |  |  |
| --- | --- | --- | --- | --- | --- | --- | --- |
|  | Proportion<br>per sample | Standard<br>deviation | Proportion<br>per sample | Standard<br>deviation | Proportion<br>per<br>sample | Standard<br>deviation | Cluster<br>marker<br>genes |
| T cells | 0.052 | 0.032 | 0.095 | 0.069 | 0.056 | 0.072 | *ETS1*<br>*CD3E* |
| Natural<br>Killer Cells | 0.008 | 0.006 | 0.025 | 0.021 | 0.01 | 0.014 | NR4A2 |
| B cells | 0.013 | 0.011 | 0.005 | 0.002 | 0.012 | 0.015 | MS4A1,<br>CD79A,<br>BANK1 |
| Plasma Cells | 0.003 | 0.004 | 0.002 | 0.002 | 0.0 | 0.0 | JCHAIN,<br>IRF4 |
| Dendritic<br>cells | 0.047 | 0.018 | 0.041 | 0.034 | 0.08 | 0.116 | CSF2RA |
| Macrophages | 0.194 | 0.124 | 0.155 | 0.145 | 0.066 | 0.069 | CD68 |
| LC1 | 0.167 | 0.071 | 0.094 | 0.04 | 0.288 | 0.12 | CLDN4,<br>CLDN3 |
| LC2 | 0.513 | 0.117 | 0.571 | 0.236 | 0.486 | 0.208 | CIDEA,<br>FOLR1,<br>NECTIN4 |
| Dividing<br>epithelial<br>cells | 0.003 | 0.002 | 0.013 | 0.013 | 0.001 | 0.001 | TUBB4B,<br>STMN1 |

We next generated a signature of genes highly expressed in MFG relative to milk cells (242 genes, MFG signature), from our bulk RNA-seq (**Fig. 3C**, **Table S2**). We used this signature to characterize which cell types could be the source of the unique RNA secreted in the MFG. We found that the MFG signature was higher in epithelial cells, and highest in the LC2 cell subset (**Fig. 3D**). In addition, the LC2 sub-population showed high expression levels for genes associated with MFG membrane formation and budding (*FABP3, BTN1A1, XDH, CIDEA*; **Fig. 3E**) (*53*) supporting the hypothesis that MFG may form primarily in LC2 cells. These results further suggest that the MFG transcriptome is more similar to LC2 cells, as compared to LC1 cells, but further studies are needed to delineate the different functions or contribution of each cell type to milk production.

### MFG bulk RNA-seq identifies transcriptional changes associated with milk production

We next generated additional bulk RNA-seq from 4, 7, and 3 selected samples collected from low, normal and high producers (respectively) during postpartum days 62-213 (mature milk) to analyze whether different levels of milk production were associated with MFG gene expression differences. Samples from low and high producers were selected to match as much as possible the days postpartum of normal producers and baby age was included as a covariate in the differential expression model (**Table 2**). Following RNA-seq, we used DESeq2 for DE analysis between milk production groups, sequentially comparing low versus normal and high versus normal samples(*51*). We found that 15 and 65 genes were downregulated between low versus normal and high versus normal, respectively; and 7 and 45 genes were upregulated between low versus normal and high versus normal, respectively (**Fig. 4A and 4B, Table S4**).

**Fig. 4:**
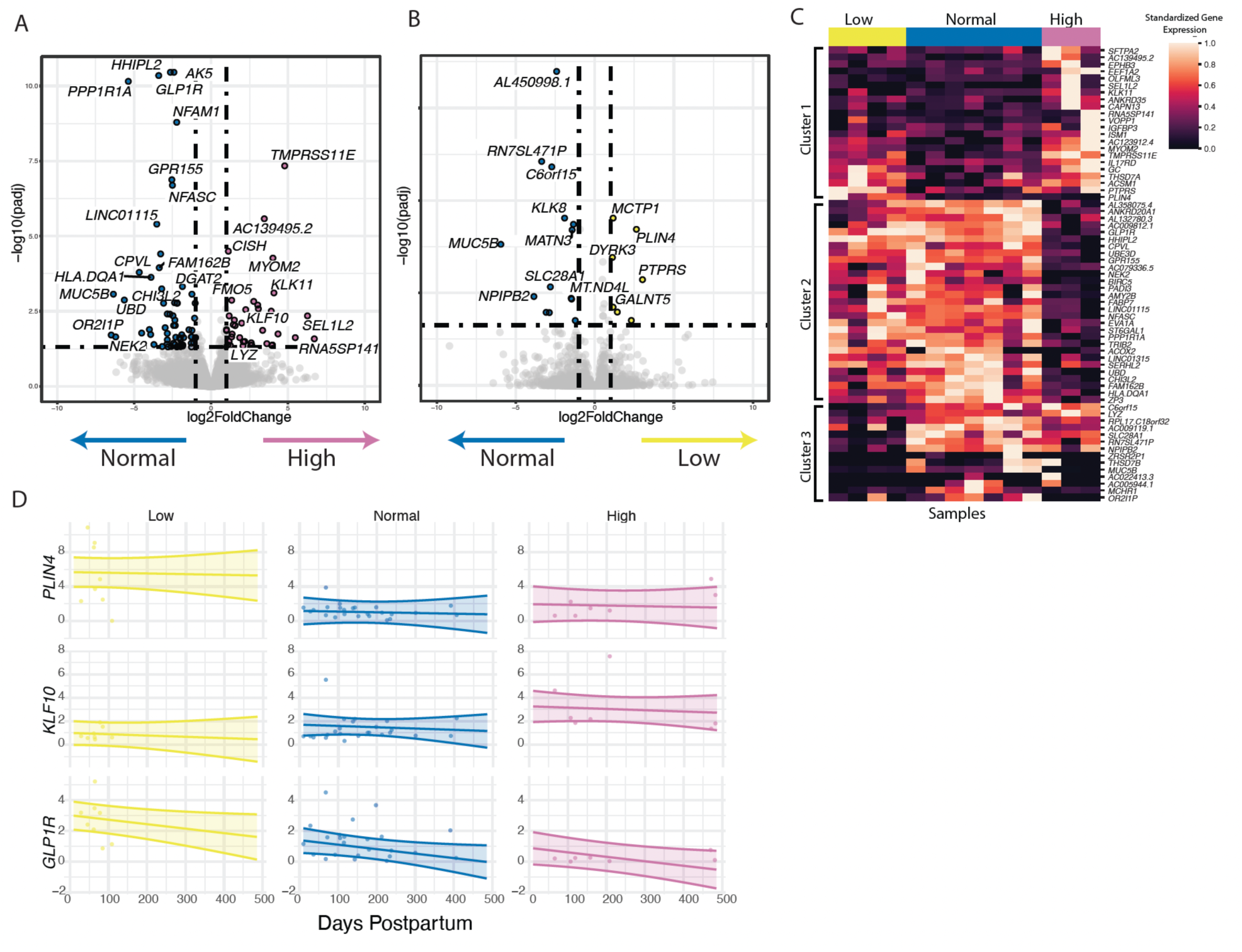
Transcriptomic changes in low and high milk production. (A) Volcano plot of DE genes between individuals with high (n=3) and normal (n=7) milk production (log-fold change logFC > 1 and BH adjusted p < 0.05). (B) Volcano plot of DE genes between individuals with low (n=4) and normal (n=7) milk production (logFC > 1 and adjusted p < 0.05). (C) Heatmap showing DE with log2foldchange > 2.5 genes between the different milk production groups colored by column-standardized gene expression. (D) qRT-PCR results for 39 MFG RNA samples at different time points postpartum. The y-axis represents the fold change in gene expression from the mean of normal producers normalized to *GAPDH* expression. A linear mixed effect model was used to determine differences in gene expression between milk production groups, controlling for days postpartum. Lines represent the fixed-effect regression with confidence intervals.

One gene that was upregulated in high compared to normal producers, *KLF10*, has been linked to the regulation of transforming growth factor beta (TGF-β) signaling(*54*); disruptions in this pathway could affect mammary gland development and lactation(*55*, *56*). *GLP1R* and *PPP1R1A* were downregulated in high compared to normal producers (**Fig. 4A**) and have been previously associated with glucose homeostasis and insulin secretion, and their dysregulation may potentially impact lactation as insulin signaling plays an important role in secretory differentiation in the mammary gland and in milk production(*15*, *57*). In addition, duodenal *GLP1R* gene expression was negatively correlated with milk production efficiency traits in dairy cattle(*58*). Currently, little is known about the role of *GLP1R* and it signaling pathway in milk production. In addition, Perilipin 4 (*PLIN4*) a gene associated with lipid accumulation in other tissues, and with breast cancer (*59*, *60*) was found in higher levels in low milk producers (**Fig. 4B**). Additional genes of interest were: cytokine inducible SH2 containing protein (*CISH*) that negatively regulates the JAK-STAT5 signaling pathway(*61*, *62*), and Lysozyme (*LYZ*) (**Fig. 4A and C**).

To validate our findings, we extracted MFG RNA from additional milk samples (total n=39 samples; 7 low, 24 normal and 8 high), and performed qPCR analysis on 5 DE genes between the milk production groups in our bulk-RNA analysis (*CISH*, *GLP1R*, *LYZ*, *KLF10*, and *PLIN4*). We calculated the fold change of each sample from the average of the normal group expression levels. We then used the mixed effect model to test if the expression of these genes is significantly different between the milk production groups, accounting for multiple samples from the same individual and the days postpartum (different times of sample collection). When controlling for time postpartum, *PLIN4* and *GLP1R* gene expression are expected to be higher in the low production group compared with normal and to high producers (p-value <0.01)(**Fig. 4D, and Table S5**). In contrast, *KLF10* gene expression is expected to be higher in high producers compared to low producers and tends to be higher compared to normal producers. We also found that *CISH* and *LYZ* expression were not significantly different between the groups over time (**Table S5 and Fig. S2A**).

### scRNA-seq identifies changes in cell type proportions that are associated with milk production and transcriptional changes in specific cell populations

We next assessed if milk production is associated with transcriptional changes in other milk cell types that are not present in the MFG transcriptome using scRNA-seq. To determine if milk cell type composition reflects milk production levels, we used our scRNA-seq data from eleven cryopreserved milk cell samples collected from our cohort (4 low, 4 normal, and 3 high production). Samples were chosen to match across time postpartum and to match the samples used for bulk RNA-seq when possible (**Table 2**). Using the nine clusters as previously shown (**Fig. 3A**), we found high variability in the composition of immune and epithelial cells between samples (**Fig. 5A**, **Table 3**); thus, we compared the composition of immune and epithelial cells separately. Using scCODA(*63*), a method for differential abundance of cell types, we found that the ratio of luminal cells 1 to 2 (LC1 to LC2) differs between samples from high production and normal production groups, with a larger proportion of LC1 cells in high production (credible effect with FDR = 0.05; **Fig. 5B**). While evaluated as credible using this analysis, the small sample size in this comparison leaves this comparison as a topic for further exploration in larger datasets. Interestingly, the low production group also exhibited a higher LC1/LC2 ratio compared to the normal group, though this difference was not statistically significant. Our analysis suggests that the proportion of LC2 relative to LC1 in milk does not directly correlate with the milk production phenotype, as both aberrant production groups displayed elevated LC1 levels. Further studies with larger sample sizes are necessary to determine whether the LC1/LC2 ratio could serve as a potential indicator of milk production dysregulation.

**Fig. 5:**
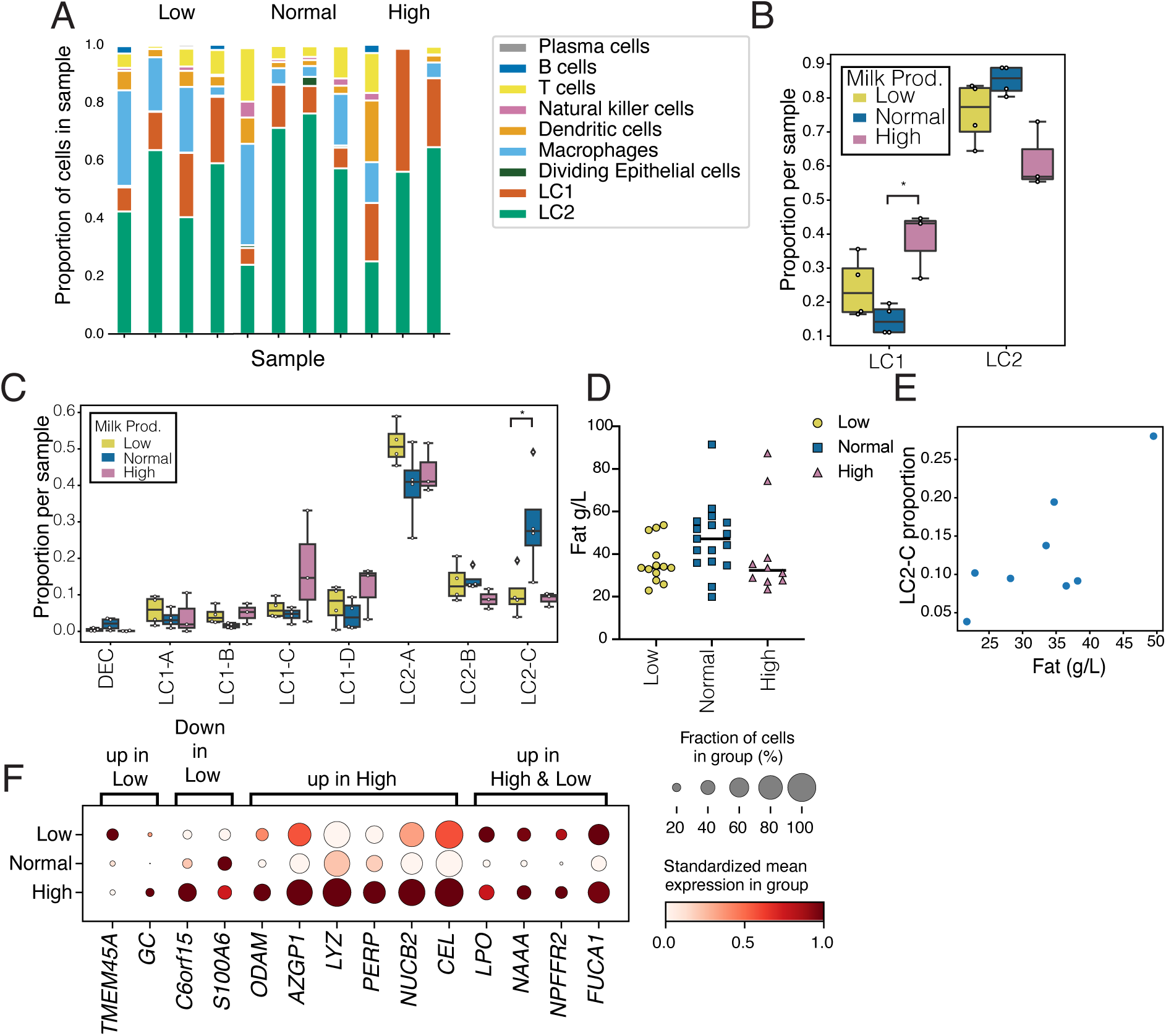
Cellular composition changes in the different milk production groups. (A) proportion of cells in each sample as revealed from scRNA-seq. (B) ratio of LC1 and LC2 subtype proportion for each sample divided into groups based on milk production. (*credible effect of difference using scCODA differential abundance) (C) Proportion of epithelial subclusters in each sample. (D) Milk fat content in samples from each milk production level. (E) Scatter plot of fat content in each sample compared to the proportion of LC2-C cells among other epithelial cells. Spearman correlation r = 0.775, p ≤ 0.03. (F) DE genes in the LC2 clustered between low, normal, and high milk producers. Dot size represents the percentage of cells in a cluster (y-axis) expressing the gene at a level > 0 and color indicates the mean log2-normalized expression of that gene in the cells in that cluster.

Among the epithelial cells, we identified eight subclusters, including four LC1 subtypes (named LC1 A-D), three LC2 subtypes (named LC2 A-C), and a small population of dividing cells (DEC) (**Fig. S3**). When comparing the proportions of these sub-populations, we found that the proportion of LC2-C cells was higher in normal suppliers compared to the low or high groups (credible effect with FDR = 0.05; **Fig. 5C**). No other changes were observed for the epithelial cell type proportions. Since the functions of the different milk cell subtypes are unknown, we decided to further characterize whether the changes in milk cell LC2-C proportions are reflected in changes in milk composition. Based on our previous finding of similarities between MFG and LC2 cells, we measured total milk fat content using the creamatocrit device (*64*, *65*) in the 40 milk samples from our cohort, including the samples used for the scRNA-seq assay. There was no significant difference in fat content (g/L) in samples provided by normal producers compared to low producers (**Fig. 5D**). Among samples with matched scRNA-seq data, fat content was positively correlated with LC2-C cell proportions (Spearman p ≤ 0.05; **Fig. 5E**). Our results identified higher proportions of LC2-C epithelial cells in normal milk suppliers, which correlated with increased milk fat content in these samples. In addition, we found that LC1-Golgi/lncRNA was negatively associated with fat content (Spearman p <0.02). No other cells types were correlated with milk fat content. To better understand the function of milk epithelial cell types, this correlative relationship between LC2-C and that LC1-Golgi/lncRNA cells and milk fat content should be further studied in future work.

Differential expression analysis between production groups within each of the epithelial cell clusters found several genes consistent with those identified in the bulk RNA-seq (**Table S6)**. For example, we identified upregulation in LC2 cells in high producers of the *LYZ* gene that encodes lysozyme, which is an anti-bacterial bioactive component found in human milk(*66*) (**Fig. 5F**; all BH-adjusted p ≤ 0.05). Several genes were increased in high suppliers in the LC2 cluster but not in the MFG transcriptome comparisons, including milk component synthesis-related genes (*AZGP1:* zinc binding and lipid metabolism; *B4GALT1*: lactose synthesis; *NUCB2*: calcium level maintenance*; KLK6:* serene protease; *GC*: vitamin D binding protein) as well as genes related to mammary gland structure (*PERP:* desmosomes; *ODAM*: epithelial cell proliferation and wound healing; *CEL:* milk synthesis related glycoprotein; **Fig. 5F**). In the same LC2 cluster, several genes were increased in both high and low suppliers compared to normal suppliers, including genes known to be related to milk synthesis (*LPO*, *NAAA, NUCA1*; **Fig. 5F**).

Immune cell subclustering identified nine clusters including three myeloid and six lymphoid cell clusters (**Fig. S4**). A majority of immune cells were myeloid cells in each sample (**Fig. S4B**). Of the myeloid cells, we identified a cluster of macrophages, dendritic cells (DC), and milk macrophages, a macrophage subset that also expresses milk production genes such as *CSN3,* and has been described in previous human milk scRNA-seq studies(*43*) and murine mammary gland studies(*67*, *68*). In the lymphoid cells, we identified clusters of natural killer (NK) cells, gamma delta (GD) T cells, CD8+ T cells, CD4+ T cells, B cells, and plasma cells using canonical marker genes (**Fig. S4**).

We next looked for differences in immune cells related to milk production, focusing on differences within myeloid cells as they are the most abundant immune cells type present in human milk. In immune cells, we found an increase in dendritic cells in individuals with high milk production compared to normal or low suppliers (credible effect with FDR = 0.05; **Fig. 6A**; **Fig. S4**). In addition, we found a trend of increased macrophage proportion in low producers. These cells also showed increased expression of genes related to TNF-alpha signaling via *NFKB* and genes related to the production of cytokines and inflammation, such as *IL1B, CCL4, CXCL3,* and *CXCL8* (**Fig. 6B, Table S6**). We scored the macrophage cells on the full TNF-alpha signaling via the NFKB hallmark pathway and observed a trend of increased gene set score in 3 out of 4 individuals with low milk production compared to normal- or high-milk producers (ANOVA p ≤ 0.1; **Fig. 6C**).

**Fig. 6:**
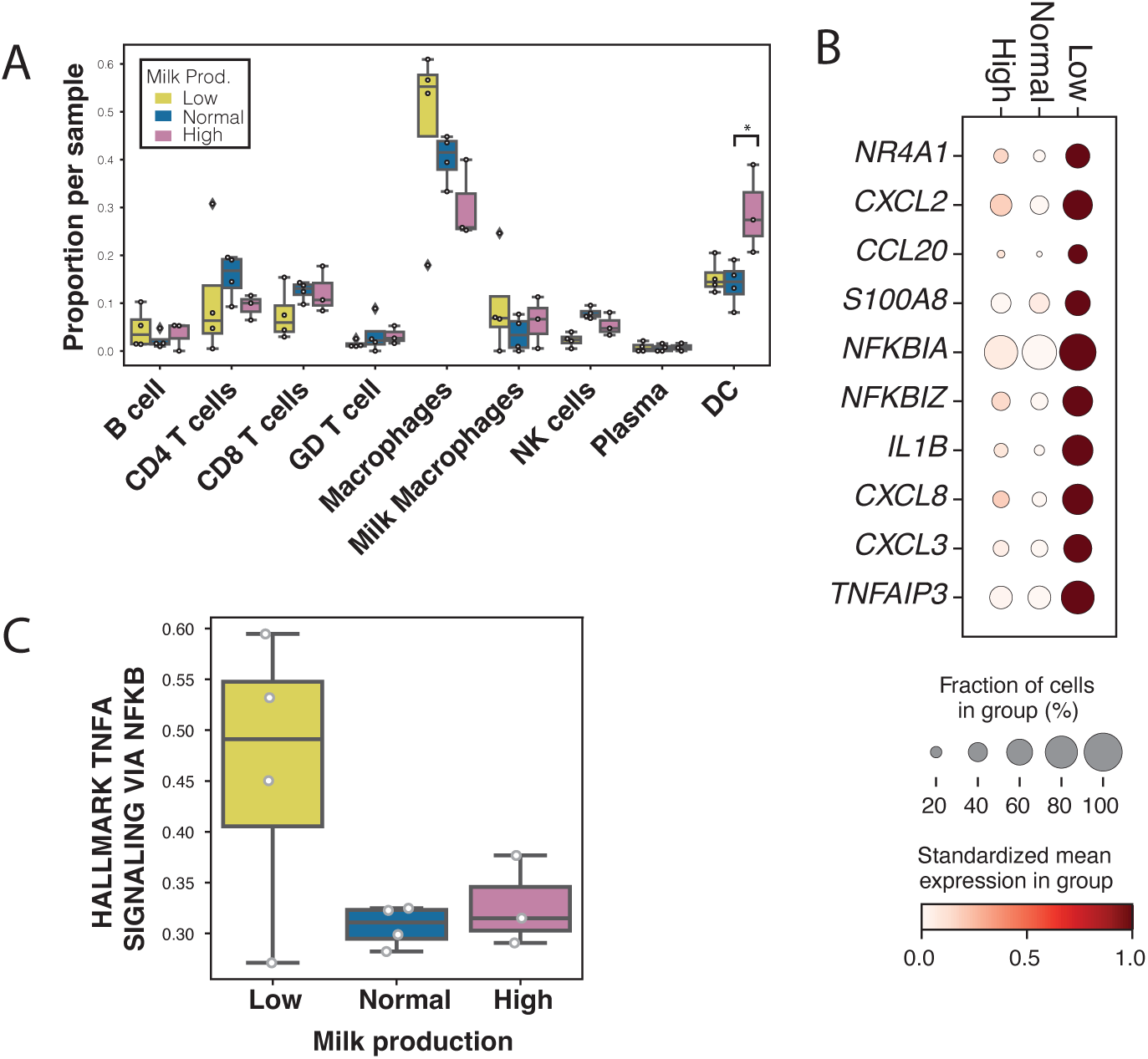
Transcriptional changes in production groups originate primarily from LC2 cells and macrophages. (A) Proportion of immune cells in each sample. (B) DE genes in the macrophages cluster between low, normal, and high milk producers. Dot size represents percentage of cells in cluster (y-axis) expressing the gene at a level > 0 and color indicates the mean log2-normalized expression of that gene in the cells in that cluster. (C) Mean gene set scores of Hallmark TNF-a signaling pathway in macrophage cells per sample in the different milk production groups computed using Scanpy gene scoring.

### Effect of milk production on the infant’s microbiome

To investigate if maternal milk production is affected by the composition of the maternal microbiome, or affects the infant gut microbiome, we collected stool samples from mothers and their infants. We analyzed 20 stool samples from 10 infants of participants with low and normal milk production and 16 samples from their 10 mothers collected at the time of milk collection. Samples were collected at multiple time points from 28 to 150 days postpartum (**Table 2**), and before the introduction of complementary foods to the infant diet. We conducted metagenomic sequencing on all samples and analyzed the data using a unique MetaPhlAn database(*69*). As only two individuals with high milk production provided stool samples, these samples were not included in the group-wise comparisons.

We next set out to characterize the infant microbiome for differences between the milk production groups. We used Principal Coordinate Analysis (PcoA) and k-means clustering (*k=3*) to characterize the dominant species in the infant gut microbial population in our cohort. We found three main clusters: 1) samples dominated by *Bifidobacterium* breve (*B. breve)*; 2) samples dominated by *Bifidobacterium longum* subsp. *infantis* (*BL. infantis)*; and 3) samples dominated by other bacteria that are typically less common in infants. Samples dominated by *B. breve* or *BL. infantis* clustered separately from samples dominated by other bacteria, revealing a distinct microbial population (**Fig. 7A**). This separation can be quantified by a lower Shannon diversity (**Fig. 7A**; point size) in the samples dominated by *B. breve* and *BL. Infantis*. One possible driver of this separation is that, when *B. breve* and *BL. infantis* are dominant, they inhibit or interfere with successful growth of other bacteria(*69*).

**Fig. 7.**
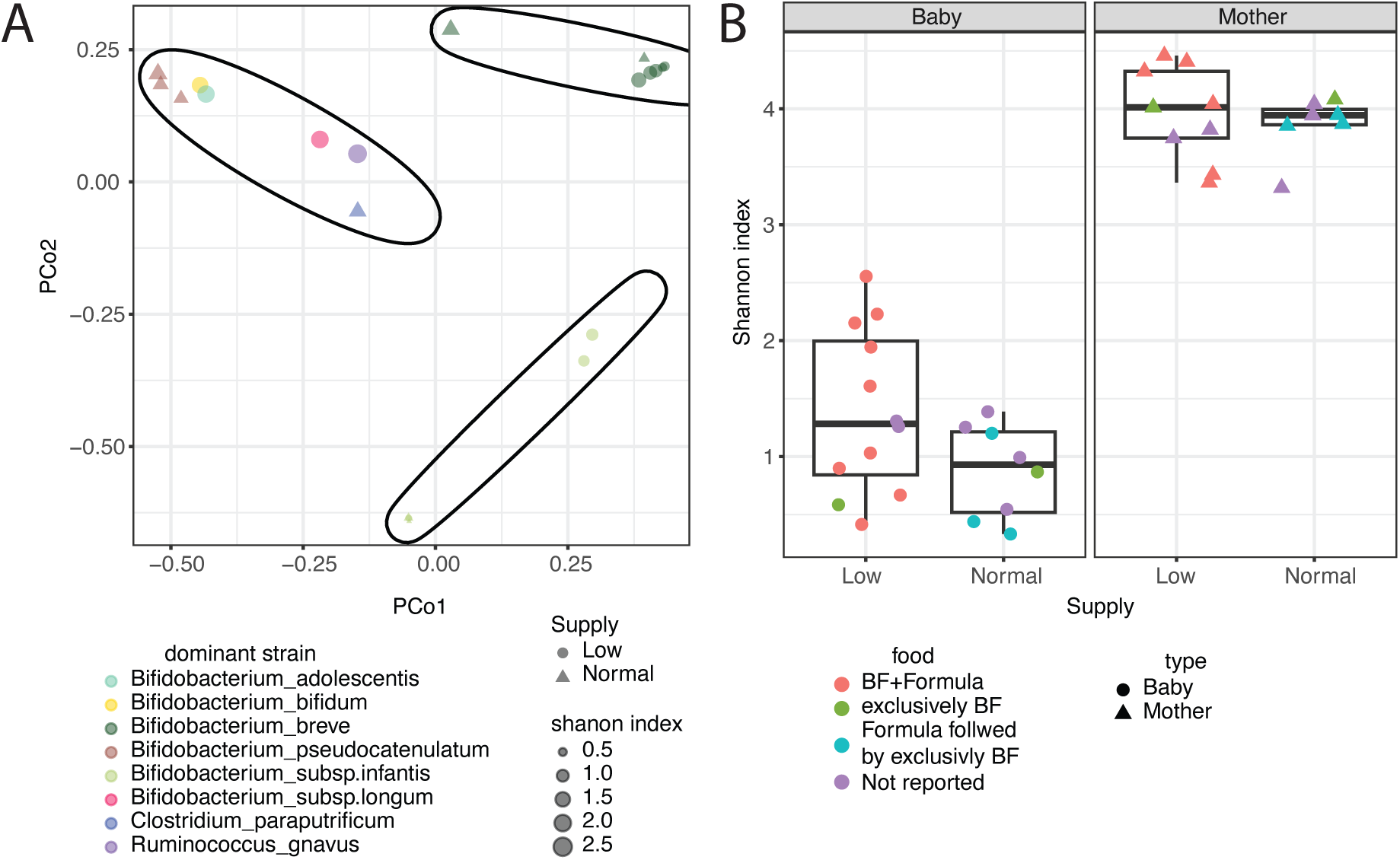
Characterization of the maternal and infant gut microbiome from different milk production groups. (A) Infant gut microbiome samples were grouped using k-means clustering with *k=3*. The samples are color-coded by their dominant bacteria, with point shapes indicating maternal milk supply (circles for low and triangles for normal milk production) and point sizes representing the Shannon diversity of each sample. (B) Shannon diversity in infant and maternal stool samples from this cohort (including only samples up to 150 days) is shown. Points are colored according to the infant feeding type at the time of sampling.

Samples from infants of mothers with low and normal supply were found in all clusters and there was no significant difference in the abundance of any bacteria species between those groups (**Fig. S5**). As expected, infant Shannon diversity index was lower than the maternal index (**Fig. 7B**). We found no difference in the maternal microbiome diversity or genus-level composition between individuals in different milk production groups in our cohort (using linear association models, while controlling for multiple samples from the same individual, **Fig. S5**). The Shannon diversity index of infants nursed by individuals with low milk production was slightly higher than that of those with normal milk production, but this difference was not statistically significant (p-value = 0.084) (**Fig. 7B**).

Since infants of low milk producers were more frequently supplemented with infant formula compared to those of normal producers (**Fig. 7B**; indicated by point color), we tested whether infant formula supplementation affected the microbiome diversity. We found that exclusively breastfed infants had lower Shannon diversity compared to those who were breastfed but also supplemented with infant formula (p-value = 0.023, **Fig. S5**). In summary, this analysis showed a trend of higher microbiome diversity in infants nursed by low compared to normal milk producers. This was likely driven by infants receiving formula supplementation (*70*), who constitute the majority in the low producers’ group, which was correlated with a more diverse microbiome compared to exclusively breastfed infants.

## Discussion

In this study, we present a comprehensive catalog of the transcriptomic and cellular changes occurring during lactation. Our findings highlight the potential of milk fat globule (MFG) RNA as a biomarker for epithelial cells, as it largely reflects the transcriptomic profile of these cells. Notably, we observed that MFG RNA contains very few unique transcripts when compared to the RNA profile of milk cells, which are commonly used as a representative of the mammary gland during lactation. This suggests that MFG RNA primarily originates from mammary epithelial cells and may serve as a useful tool for studying epithelial-specific gene expression in lactation research. Our analysis also revealed that MFG RNA is not enriched in immune-related transcripts, indicating a low contribution from immune cells. This distinction is particularly important for studies focusing on immune function during lactation, as MFG RNA does not provide a reliable representation of immune cell activity. Instead, researchers investigating immunological aspects of lactation may find milk cells to be a more suitable source of RNA.

In addition, this study is the first to analyze changes in transcriptomic in MFG and milk cells collected from fresh milk samples from individuals with low, normal, and high human milk production. Previous studies that examined changes in the milk cell transcriptome of low milk producers were limited by confounding from a higher body mass index (BMI) among individuals with low milk production compared to the normal production group(*13*, *14*). High BMI may lead to greater changes in milk production that are dependent on systemic inflammatory state as well as local inflammation in the mammary gland. We observed a trend of increased TNF-a signaling in macrophage cells in low producers in our scRNA-seq analysis, but this pathway was not different in our bulk RNA-seq analysis in MFG. In our cohort, there was no significant difference in BMI between the study groups, which may explain the smaller differences in the milk inflammatory markers between our study groups compared to previous studies (*71*). Another study that used frozen milk samples from eight participants found no differences in gene expression between individuals with low and high milk production (*72*). This might be due to the use of frozen milk samples, that is cause fast degradation of RNA and reduce significantly the RNA integrity (RIN score). The same study reported that individuals with low milk production (*n=4*) have more depression and anxiety compared to individuals with high production (*n=4*) (*72*); however, depression did not differ between the groups in our study and we did not assess anxiety. Since breastfeeding is a multifactorial process that depends on many environmental and internal factors(*73*), it will be essential in the future to quantify differences in these confounders within and between the study groups.

Our analysis provides important insights into changes in the transcriptional signature and cell type composition of the mammary gland under aberrant milk production. We found that most genes that are required for milk production, including milk proteins (caseins and lactalbumin) and milk fat secretion, are not differentially expressed between the milk production groups, and that the conserved cellular milk production machinery functions equivalently across all groups. Our findings also identify gene targets that should be further studied to understand their role in human milk production.

Our study is the first to our knowledge to examine milk cell population proportions in samples collected from individuals with low, normal, and high milk production. In terms of total cell proportion, we did not observe any significant differences between our study groups, possibly due to the high variability of immune cells in milk samples and the small sample size in our study. Focusing on epithelial cells alone, the LC2-C subcluster (high *KRT*) was depleted in low and high milk production groups. Along with the correlation between milk fat concentrations and LC2-C cells percentages, these results suggest a role for LC2-C cells in the fat content in milk and should be further characterized. In immune cells, dendritic cells (DC) were increased in the high production group. Interestingly, the DC population was previously shown to increase during involution in mice(*74*); our results suggest that high milk production might lead to similar cellular changes as in involution due to the constant attempt to reduce milk production in these individuals.

Individuals with low milk production are usually using breast milk pumps to try and increase their milk supply and are often recommended to add infant formula to their infant diet. These different feeding behaviors might also affect the infant microbiome(*75*), and we therefore aimed to identify if impaired milk production is associated with changes in the infant microbiome. Our findings revealed no differences in microbiome diversity or species composition between mothers and infants with low versus normal milk production. Our results align with earlier studies from larger cohorts that demonstrated that combining formula feeding with breastfeeding tends to increase microbial diversity compared to exclusive breastfeeding; however, partial and exclusively breastfeeding reduces microbial diversity compared to exclusively formula feeding (*76*). These findings further support the messaging that individuals with low milk supply should be encouraged to continue partial breastfeeding to support healthy infant microbiome development.

Our study has some important limitations. This is a small prospective study, and samples were collected at a relatively late stage in lactation due to the need for lactation consultants to study lactation in each participant prior to recruitment. Furthermore, larger studies that follow up on lactation in the participants from childbirth through the mature milk stage are needed to better understand the changes occurring just after birth and their effect on establishing lactation and breastfeeding outcomes. In addition to the changes detected in the milk cell populations, it is possible that there are other changes in cell populations that are not shedding into milk (basal cells, stromal cells)(*44*), which affect the ability individuals to produce or secrete milk. Moreover, hormonal regulation and other upstream factors may also play a role in milk production(*62*); these factors were not examined in our analysis and cannot be detected using milk cells. Our microbiome data is limited because it does not contain samples of individuals with high milk production, and by the small sample size of the low and normal producers.

In summary, our study provides a unique database of gene expression during lactation and that can be used to find specific genes that might promote or inhibit milk production in cases of aberrant milk production. However, the specific role that these genes play in human milk production needs further research. Our study will pave the way for more research in the area of milk production using genomic characterization, and future studies will assist in our understanding, diagnosis, and treatment of breastfeeding difficulties.

## Materials and Methods

### Cohort

#### Participant cohort and data collection

milk samples for this study were collected in two independent cohorts and the institutional review board of the University of California, San Francisco, approved these studies (UCSF Milk production study #19-29297 and COVID-19 Vaccine in Pregnancy and Lactation (COVIPAL) cohort study #20-32077). Written informed consent was obtained from all study volunteers. The COVIPAL samples were collected and stored as previously described(*77*) and were used as additional samples for fat layer RNA-seq.

### Clinical data

#### Data collection

Data was collected through an online questionnaire that was sent to participants via email using REDCap. To assess perceived insufficient milk supply (PIMS), mothers in both cohorts were asked to report if they feel that they produce too much/normal/not enough breast milk to meet their infant’s needs. In the UCSF Milk supply study, mothers were also assessed by lactation consultants to address any obvious reason for low/high milk production and were also asked whether their infants are satisfied with the volume of milk they produced.

#### Statistical analysis

Surveys data was exported from RedCap and analyzed on R version 4.3.2 (2023-10-31) using the CompareGroups(*78*) function to generate Table 1.

### Milk sample collection and processing

Fresh human milk samples were self-collected by participants into sterile containers using Ameda Dual HygieniKit Milk Collection System provided to the participants. For each collection, mothers were instructed to use a new/autoclaved sterile kit, wash hands before pumping and empty both breasts. Ten to thirty milliliters of milk were collected from each breast for analysis. Milk from each breast were processed separately. Mothers that prefer using different pumps were asked to sterilize the pump parts at home and document it. Samples were transported on ice from the participant’s home to the lab for processing. Milk samples were collected and processed as soon as possible and no longer than 2 hours after expression by the study staff. In the lab, whole milk was aliquoted, and the rest was centrifuge at 800g for 20 minutes in 4°C to pellet the cells and separate the fat layer. Fat layer was removed using a sterile spoon and mixed with RLT lysis buffer or Trizol for RNA extraction using the RNeasy kit (Qiagen). Supernatant was aspirated and aliquoted and milk cells pellet was resuspended in PBS with 0.04-1% bovine serum albumin (BSA). Cells were washed once or twice and were analyzed immediately or cryopreserved using CryoStor CS10 freezing media (STEMCELL Technologies). For milk fat content, frozen milk samples were thawed to room temperature and were gently mixed to homogenize before measured. Milk fat content was measured using Creamatocrit Plus (EKF Diagnostics). On outlier sample that had fat levels above the normal range expected for mature human milk (8 g/dL)(*79*) was excluded from further analysis related to fat composition.

### Bulk RNA-seq

Total RNA samples from milk fat layer of milk cells were sequence at Novogene Co. ltd. Libraries were generated using the SMARTetr V2 or V3 kits. The sequencing was performed using a paired end 150bp strategy on the Illumina platform (Illumina NovaSeq 6000). Sequencing reads were aligned to reference human genome hg38 using STAR aligner. Normal breast RNA-seq data was downloaded from GEO (PRJNA292118)(*49*). Read counts were normalized to transcripts per kilobase million (TPM). Quality control matrixes and details about sequence reads/samples are in **Table S8**.

### Bulk differential analysis

Differential expression between paired milk cell and fat layer transcriptional data was performed using DESeq2 (*51*) with designs ‘~ subject+layer’ for milk cells vs fat layer, ‘~tissue_origin’ for cells or fat vs breast tissue, and ‘~baby age + milk_supply’ for milk supply. For milk cells vs milk fat, genes with adjusted pvalue less than 0.01 and log2foldchange greater than 2 or less than −2 were included in enrichment analyses. For milk supply comparisons, genes with adjusted pvalue less than 0.05 and log2foldchange greater than 1 or less than −1 were included in enrichment analyses.

### Bulk gene set enrichment analyses

Gene ontology enrichment was run using the clusterProfiler package in R with function enrichGO using biological processes ontologies with Benjamini-Hochberg p-value correction for multiple tests and genes expressed in dataset as background followed by the simplify function to remove redundant gene ontology hits(*48*).

### qPCR for gene expression-

MFG were preserve in RLT-buffer or RNAlater and used for RNA isolation using RNeasy kit (Qiagen,#74134). Reverse transcription was performed with 1μg of total RNA using qScript cDNA Synthesis Kit (Quantabio, 95047) following the manufacturer’s protocol. qRT-PCR was performed on QuantStudio 6 Real Time PCR system (ThermoFisher) using PerfeCTa™ SYBR® Green FastMix™, Low ROX™ (QuantaBio, 95074). Statistical analysis was performed using ddct method with *Gapdh* primers as control (see primer sequences in **Table S7**). Gene expression results were generated using mean of all participants with normal milk production as the reference value.

Linear mixed effect model was generated in R using the lmerTest package and the following formula: gene, “~ Supply_group + Days_pp + (1 | ‘Study ID’).

The emmeans function was used for post-hoc analysis to compare the means of the difference milk production groups using Tukey methods for comparing a family of 3 estimates, with Degree-of-freedom method: Kenward-roger (results in **Table 5S**).

### Single cell RNA-seq

Cryopreserved cells were thawed and washed with 10 ml of Mammary Epithelial Cell Growth Medium (PormoCell) and later with 2 ml of PBS+1-2% BSA. Live cells were counted using Acridine orange (AO)/propidium Iodine (PI) staining using DeNovix Celldrop cell counter. Twenty-five thousand cells from each sample were loaded on the 10X single cell chip. Chromium Next GEM Single Cell 3ʹ Kit v3.1 was used following the manufacturers’ protocol.

### Single cell RNA-seq analysis

#### Alignment

Samples were aligned to hg38 using 10x Genomics CellRanger v6.1.2.

#### Preprocessing

Using the Scanpy package v1.9.6, data was filtered to remove cells with fewer than 200 genes and genes expressed in fewer than 10 genes(*80*). Expression was normalized to 1e4 total counts per cell and the log base 2 + 1 was taken.

#### Clustering analysis

Following a standard Scanpy pipeline (*80*), highly variable genes were selected using the scanpy highly_variable_genes function with batch key=sample. Twenty-six principal components and ten neighbors were used to construct the neighborhood graph. Clustering was performed iteratively beginning with identification of broad immune and epithelial cell clusters using Leiden clustering followed by re-selection of variable genes on epithelial and immune subsets then followed by further subclustering on each broad celltype: T cells, B cells, myeloid cells, LC1s, and LC2s. In each subclustering stage, clusters were identified as doublet cells and removed if they contained marker genes from distinct lineages. During subclustering, Harmony was used for batch integration(*81*). Celltype labeling was done based on comparison of marker genes to those identified in prior human milk cell datasets(*42–44*).

#### Differential expression

To define marker genes for each subcluster as genes which identify cells in that cluster regardless of their sample of origin as well as genes specific to supply level groups which are reproducible across individual samples, we used pseudobulk differential expression analysis as previously described(*43*, *82–84*). Briefly, we generated pseudobulk counts for the cells in each sub-cluster in each sample by summing the raw counts for each group and used these to identify differentially expressed genes between subclusters and between conditions in a sample aware manner. We removed subcluster/sample pools with fewer than 10 cells from these comparisons. Using the DESeq2 package (*51*), we performed differential expression analyses using the Wald statistical test with the design formula “~ donor + celltype” to identify celltype marker genes and “~ donor + baby age + Milk supply” to identify milk supply differential genes for each cluster(*51*). Marker genes were then filtered for adjusted p value < 0.05, log2foldchange > 0.4 and proportion of cells in cluster expressing the gene > 0.4. Milk supply level differential genes were filtered for adjusted p value < 0.05 and proportion of cells in cluster expressing the gene > .1 full differential gene lists are available in **Tables S1-4**.

#### Gene set enrichment analysis

Functional enrichment analysis on these differential genes was performed using Enrichr using the gseapy package with the gene set GO_Biological_Processes_2023(*85*, *86*). For the macrophage cluster, TNFA signaling via NFKB score was calculated using the full list of genes with the scanpy function “score_genes”.

#### Statistical analysis of scRNA-seq results

The scCODA package was used to test statistically differentially abundant celltype proportions in the scRNA-seq data between low, normal, and high supply groups(*63*). This approach compares the ratio of celltype abundance within a sample to a reference celltype. Since this study did not include a clear cell type whose abundance should be expected to stay stable, the scCODA test was run iteratively with each cell type as the reference cell type, and a cell type was considered differentially abundant if the scCODA test identified it as significantly differential with more than half of the other cell types as the reference. This test was run on the cell type composition across all cell clusters as well as within just the immune cells and just the epithelial cell subclusters separately.

All RNA quality control matrixes for the bulk and scRNA sequence analysis are in **Table S8**.

### Stool samples collection

Maternal and infant stool samples were collected on the day before, on the day of, or the day after milk sample collection. Maternal stool samples were collected using a Feces Catcher (Zymo Research, R1101-1-10) following the manufacturer’s instructions, then immediately scooped into DNA/RNA Shield fecal collection tubes (Zymo Research, R1101-E). Infant stool samples were collected directly from the diaper (with mothers instructed not to apply cream at the time of sample collection) immediately after bowel movement into DNA/RNA Shield fecal collection tubes (Zymo Research, R1101-E). Samples were brought to the lab within 3 hours of collection or kept frozen in a household freezer if collection was scheduled for a later time point. Samples were shipped to the lab on ice and stored at –80°C until analysis.

### DNA Extraction

Stool samples underwent DNA extraction performed by the Microbial Genomics Core at the University of California, San Francisco, using a modified cetyltrimethylammonium bromide (CTAB) buffer-based protocol as described in a published article (*87*). Briefly, each frozen stool pellet was suspended in 500 µL of 5% CTAB extraction buffer in a Lysing Matrix E tube (MP Biomedicals) by vortex and incubating at 65 °C for 15 minutes. Then, 500 µL of phenol:chloroform:isoamyl alcohol (25:24:1) was added, followed by bead-beating at 5.5 m/s for 30 seconds and centrifugation at 16,000g for 5 minutes at 4 °C. The aqueous phase (~400 µL) was transferred to a new 2mL Eppendorf tube, and the extraction was repeated with an additional 400 µL of 5% CTAB buffer, yielding ~800 µL from repeated extractions. Chloroform was then added in equal volume, mixed, and centrifuged (16,000g for 5 minutes). The resulting aqueous phase (~500 µL) was combined with 2 volumes of 30% polyethylene glycol (PEG)/NaCl solution and stored at 4 °C overnight to precipitate DNA. Samples were then centrifuged (3,000g for 60 minutes), washed twice with ice-cold 70% ethanol, and resuspended in 100 µL of sterile water. DNA from each sample was quantified using the Qubit 2.0 Fluorometer with the dsDNA BR Assay Kit (Life Technologies, #Q32853).

### Shotgun Metagenomic Library Preparation

Shotgun metagenomic DNA library preparation was performed using the Illumina DNA Prep Kit (Illumina, #20060059) according to the manufacturer’s instructions. A total of 150 ng of input DNA from each sample was used for library preparation, which involved enzymatic fragmentation (tagmentation), index-adapter ligation, and amplification. The Illumina libraries were quantified using the Qubit 2.0 Fluorometer with the dsDNA High Sensitivity Assay Kit (Life Technologies, #Q32854) and pooled at equal molar concentrations. The final pooled libraries were submitted for sequencing at the Center for Advanced Technology (CAT) at UCSF, where they were sequenced using the Illumina NovaSeq 6000 in a 2×150 bp paired-end run protocol.

### Metagenomic analysis

Host reads were removed using an in-house pipeline by aligning reads to the human genome by Bowtie2(*88*) (2.4.5-1). Samples were filtered and trimmed for Nextera adaptors using fastq-mcf, ea-utils(*89*) (1.05). Taxonomic profiling was done using MetaPhlAn4(*90*) with a custom database which allows quantification of *B. longum* subspecies(*69*). Further analysis was done using an in-house R (4.2.2) script utilizing dplyr(*91*) (1.1.2), tidyr(*92*) (1.3.0), tidyverse(*93*) (2.0.0). Plots were created using ggplot2(*94*) (3.4.2) colors were used from RColorBrewer(*95*) (1.1-3) and pals(*96*) (1.7). Alpha and beta diversity were calculated using “diversity” (Shannon index) and “vegdist” (Bray-Curtis dissimilarity) from the vegan(*97*) (2.6-4) package and the PcoA was created using the ape(*98*) (5.7-1) package. Independent t-test test was performed to test between groups when mentioned using the R function “t.test”. In addition, the “Maaslin2”(*99*) R package was used to perform linear models in order to find associations between breast milk supply and species in the infant gut. The individual was set as a random factor to account for the effect of each mother-infant pair.

## Data Availability

All data produced in the present study are available upon reasonable request to the authors

https://www.ncbi.nlm.nih.gov/geo/query/acc.cgi?acc=GSE275626

## Acknowledgments

We thank the donors and their families for participation in this study. We thank the lactation consultants at UCSF clinic for supporting this work. This work is supported by funding from The Benioff Center for Microbiome Medicine at UCSF. We thank the UCSF Benioff Center for Microbiome Medicine Microbial Genomics CoLab Plug-in for their assistance in data generation. We thank Matt Thomas and the Cornell Statistical Consulting Unit for their assistance with statistical analysis.

## Funding

These studies were supported by the Weizmann Institute of Science - National Postdoctoral Award Program for Advancing Women in Science, the International Society for Research in Human Milk and Lactation (ISRHML) Trainee Bridge Fund, the Human Frontier Science Program and the National Institute of Child Health and Human Development (NICHD) 1K99HD113880 (to YG). National Institute of Allergy and Immunology K08AI141728 and a generous gift from the Krzyzewski Family (to SG). NIH - National Human Genome Research Institute R01 HG012967 and NIH National Cancer Institute 5U2CCA233195 (to BEE), and BEE is a CIFAR Fellow in the Multiscale Human Program. NIH F32 (F32 HD114427) (to SKN). Benioff Center for Microbiome Medicine UCSF (to NA).

## Author contributions

Conceptualization: YG, NA, VJF

Methodology: YG, SKN, JZ,

Clinical studies design, supervision and samples collection: YG, MP, SLG, VJF

Investigation: YG, SKN, ZL, DE, EB, ARK

Visualization: SKN, DE, YG

Supervision: NA, VJF, BEE, MY

Writing—original draft: YG, SKN

Writing—review & editing: NA, VJF, BEE, MY, MP, SLG

## Competing interests

BEE is on the Scientific Advisory Board for ArrePath Inc, Crayon Bio, and Freenome; she consults for Neumora. SKN reports compensation for consulting services with Radera Biosciences. All other authors declare that they have no competing interests.

## Data and materials availability

All data are available in the main text or the supplementary materials. Sequencing data is available at:

1. https://www.ncbi.nlm.nih.gov/geo/query/acc.cgi?acc=GSE275626
2. https://www.ncbi.nlm.nih.gov/geo/query/acc.cgi?acc=GSE275627

## Notes

### Author Declarations

The institutional review board of the University of California, San Francisco, gave ethical approval for this work. studies (UCSF Milk production study #19-29297 and COVID-19 Vaccine in Pregnancy and Lactation (COVIPAL) cohort study #20-32077)

