## Supplementary Materials for "Genomic characterization of normal and aberrant human milk production"

Yarden Golan *et al.*

**This PDF file includes:**

Figs. S1 to S5

Tables S1 to S8

Fig. S1. Cellular composition of human milk cells. (A) proportion of each cell type found in human milk. Each dot represents a different sample (n=11). (B) Marker genes expression for each cluster of human milk cell type (x-axis). Dot size represents percentage of cells in cluster (y-axis) expressing the gene at a level > 0 and color indicates the mean log2-normalized expression of that gene in the cells in that cluster.

Fig. S2. qPCR of genes not significantly different between groups and Milk Cell signature on scRNA-seq clusters. (A) qRT-PCR results for 39 MFG RNA samples at different time points postpartum. The y-axis represents the fold change in gene expression from the mean of normal producers normalized to GAPDH expression. A linear mixed effect model was used to determine differences in gene expression between milk production groups controlling for days postpartum. Lines represent the fixed-effects regression line with confidence intervals. (B) Gene expression scores for scRNA-seq cell clusters with a signature generated by differential expression between bulk RNA-seq on milk cells compared to the milk fat layer RNA-seq (shown in Figure 3C in red).

Fig. S3. Epithelial cells sub-clustering. (A) UMAP low dimensional visualization of scRNA-seq data of epithelial clusters colored by sub-type cluster. (B) LC1 and LC2 subtype proportion for each sample (x-axis) divided into groups based on milk production. (C) Marker genes for epithelial cells sub-clusters. Dot size represents percentage of cells in cluster (y-axis) expressing the gene at a level > 0 and color indicates the mean log2-normalized expression of that gene in the cells in that cluster.

Fig. S4. Immune cells sub-clustering. (A) UMAP low dimensional visualization of scRNA-seq data of immune cells clusters colored by sub-type cluster. (B) Marker genes for immune cells sub-clusters. Dot size represents percentage of cells in cluster (y-axis) expressing the gene at a level > 0 and color indicates the mean log2-normalized expression of that gene in the cells in that cluster.

Fig. S5. Abundance of main bacterium types and effect of formula supplementation on Shannon diversity. (A) Bar plot of infant microbiome at the species level. 20 samples from 10 infants up to age 150 days were analyzed. The main bacteria types are shown and the rest are categorized as “Other”. (B) Bar plot of maternal microbiome at the genus level. The main bacteria are shown and the rest are categorized as “Other”. (C) Shannon diversity in infant stool samples from this cohort (including only samples up to 150 days). Points are colored according to the infant feeding type at the time of sampling. There was significant difference in Shannon diversity between the feeding types (p-value = 0.023).

**Supplementary tables:**

**Table S1: Additional information on formula supplementation and lactation consultant summary for each participant.** Information was extracted from electronic medical records.

**Table S2: Differentially expressed genes and pathways between milk cells, milk fat globules and breast tissue.**

**Table S3: Marker genes for general celltypes.** Gene lists identified using pesudobulk DESeq2 analysis of celltype vs other celltypes filtered for adjusted p values < 0.05, Log2foldchange > 0.4 and percent celltype expressing >0.3.

**Table S4: Differential expression and GO biological process annotation results on bulk RNA seq data from fat layer samples comparing milk production levels using DESeq2.**

**Table S5:** **Milk production groups comparisons for qPCR gene expression patterns.** Results from the post-hoc emeans analysis that compare between the 3 groups of milk production tested in this study.

**Table S6: DE genes between milk production groups in different the cells types based on scRNA expression.**

**Table S7: qPCR primers**

**Table S8: quality control matrixes for bulk and sc RNA sequences.**
